# A comparison of core temperature at different sites during cytoreductive surgery with hyperthermic intraperitoneal chemotherapy: A prospective observational study

**DOI:** 10.1101/2024.11.27.24318051

**Authors:** Jung–Won Kim, Jin Ho Kim, Soojeong Oh, So Woon Ahn

## Abstract

**Background:** During cytoreductive surgery with hyperthermic intraperitoneal chemotherapy, body temperature rapidly changes due to the hyperthermic agent. Depending on the method used, obtained values may vary, particularly during localized heating of the abdominal cavity. The primary aim of our study was to compare the effectiveness of core temperature monitoring techniques, nasopharyngeal temperature, zero-heat flux cutaneous thermometer device, and esophageal temperature, during hyperthermic intraperitoneal chemotherapy with cytoreductive surgery.

**Methods:** Body temperature was measured using a zero-heat flux cutaneous thermometer device on the forehead and esophageal and nasopharyngeal probes throughout the surgery. Temperature differences were then calculated for each pair of measurements. We conducted an agreement analysis between the nasopharyngeal temperature and reference core temperature measurements using the 95% Bland–Altman limits of agreement for repeated measurement data. The proportion of all differences that were within 0.5 °C and repeated measures Lin’s concordance correlation coefficient were estimated.

**Results:** The mean overall difference between nasopharyngeal and forehead temperature was -0.04 ± 0.33 °C (95% limits of agreement: -0.69–0.62), and esophageal temperature was 0.02 ± 0.35°C (95% limits of agreement: -0.66–0.71). The proportion of differences within 0.5 °C of TSpotOn was 0.03 (95% CI 0.01–0.04) and that of esophageal temperature was -0.06 (95% CI -0.08 to -0.05). The Lin’s concordance correlation coefficient of both were 0.94 (95% CI 0.93–0.94). After the hyperthermic intraperitoneal chemotherapy period, the temperatures rapidly decreased, followed by a noticeable rebound increase, particularly nasopharyngeal and forehead temperatures. The maximum difference between esophageal temperature and other temperatures was 1.5 °C.

**Conclusions:** Our results reveal a strong correlation between nasopharyngeal, zero-heat flux cutaneous, and esophageal temperatures during surgery. However, consistent temperature differences with the esophageal thermometer were not observed. Post-hyperthermic intraperitoneal chemotherapy, rebound heating occurred at all sites, notably in the nasopharyngeal and zero-heat flux cutaneous areas.

## Introduction

Hyperthermic intraperitoneal chemotherapy (HIPEC) has emerged as a promising therapeutic modality for patients with peritoneal or metastatic cancer in the abdominal cavity [1], demonstrating significant improvements in survival rates compared with traditional palliative approaches. The high-concentration delivery of heated intraperitoneal chemotherapy, with limited systemic absorption [2], has transformed the management of these patients. Meticulous and vigilant care is essential for anesthesiologists during HIPEC, with or without cytoreductive surgery, including managing blood loss, fluid shift, hypothermia, and hemodynamic changes induced by hyperthermia.

Elevations in body temperature lead to physiological changes that manifest as a gradual rise in heart rate and end-tidal carbon dioxide values, accompanied by metabolic acidosis and elevated arterial lactate values [3,4]. The central nervous system, among the major organ systems, is particularly susceptible to hyperthermia [5]. Studies on induced hyperthermia in healthy volunteers showed impaired memory at a core temperature of just 38.8°C compared to normothermia [6].

Under normal conditions, the body temperature is not uniform, typically exhibiting a 2–4 °C difference between core and shell temperatures. Vasomotor thermoregulation is activated in response to environmental temperature changes, facilitating heat redistribution to maintain a stable core temperature. However, variations in core temperatures can occur during rapid cooling or rewarming processes [7].

During extensive debulking procedures, body temperature commonly decreases; however, it rapidly increases with the administration of a hyperthermic (42–43 °C) cytostatic agent [8–10]. Accurate and appropriate temperature monitoring is crucial for maintaining normothermia throughout the procedure.

Depending on the method used, obtained values may vary, particularly during localized heating of the abdominal cavity. Although the gold standard for core temperature monitoring is the pulmonary arterial catheter, its invasiveness necessitates the use of alternative methods such as esophageal and nasopharyngeal probes and zero-heat flux cutaneous thermometers (ZHF) [11–13].

The primary aim of our study was to investigate and compare the effectiveness of nasopharyngeal temperature monitoring with two core temperature measurement techniques, the ZHF device and esophageal temperature, during HIPEC with cytoreductive surgery.

## Materials and methods

This study received approval from the Ethics Committee for Research with Medicine of CHA Bundang Medical Center, CHA University Hospital, Gyeonggi–do, Korea on August 5^th^, 2019 (approval number: CHAMC–04–050; registration number: KCT0003980). Written informed consent was obtained from all participants.

Data were collected from July 04, 2019, to Dec 22, 2022. During this period, 55 patients, who had previously signed a written informed consent form, underwent cytoreductive surgery with HIPEC at our hospital.

We excluded patients with systemic or local infections of the forehead or any contra-indication to esophageal and nasal probe insertion, as well as those who changed their chemotherapy plan for several reasons. Finally, 41 patients participated in the study. (Fig 1)

**Fig 1.**
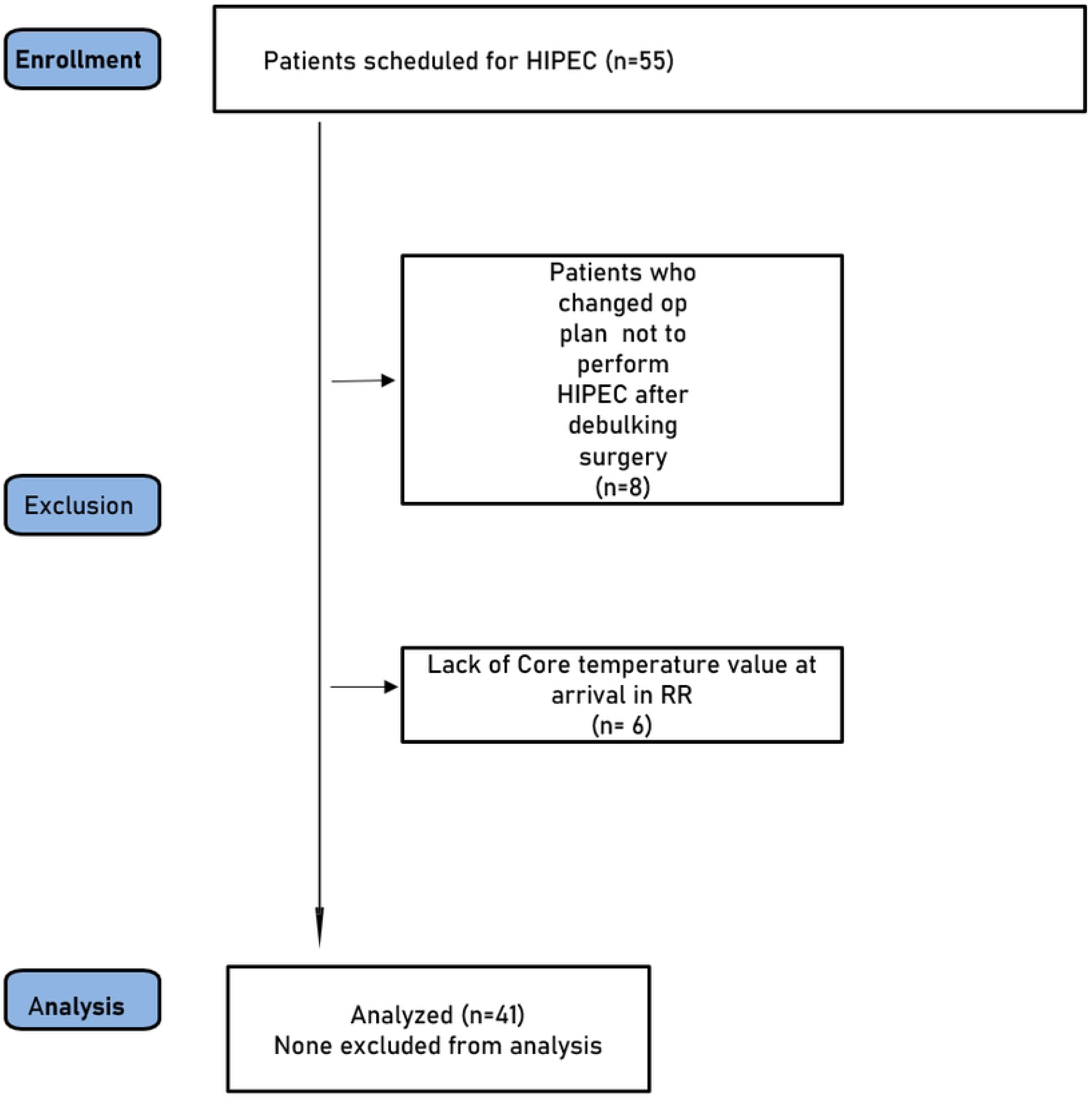
Flow chart

Uniform balanced anesthetic techniques and volatile agents were used in all patients. Following anesthesia induction, temperatures were measured at various sites, as follows: on the forehead with the ZHF, via a SpotOn® sensor (3M™ Bair Hugger™ Temperature Monitoring System, 3M, St Paul, MN, USA); at the nasopharynx via a probe (E-temp, Ewha Biomedics, Seoul, Korea) inserted to a depth equal to the distance between the nares and the earlobe and at a distance of 30 cm from the nostrils or 20 cm from the lips; and with an esophageal probe (E-temp, Ewha Biomedics, Seoul, Korea), placed just after tracheal intubation (following the Mekjavić and Rempel formula [14]). Temperature measurements were simultaneously collected at 5-minute intervals using each device throughout the surgery. Temperature differences were then calculated for each pair of measurements.

### Statistical analysis

A Bland–Altman analysis, which incorporated multiple observations per individual was used to evaluate the comparability of the temperature readings between nasopharyngeal temperature (Tnaso) and other measurement sites [15]. If the observed limits of agreement (± 1.96 SD around the mean difference) were clinically accepted, indicating that 95% of the differences were expected to fall within these limits, the two methods were considered equivalent. A priori, the acceptable limits of agreement were set at ≤ 0.5 °C, a clinically relevant threshold that is based on the usual temperature cycle variations in human beings and is associated with clinical complications [16]. Additionally, a Bland–Altman plot illustrating individual differences between the two measurements versus their average was generated to show agreement with the reference method (the smaller the range between these two limits, the better the agreement). We calculated the proportion of the Tnaso measurements that were within 0.5 °C of the corresponding method of reference, and the 95% confidence interval for the proportion was estimated using bootstrap percentiles based on 10,000 resamples. To assess reproducibility, Lin’s concordance correlation coefficient (LCCC) was computed and interpreted using McBridés strength-of-agreement criteria for continuous variables (almost perfect: > 0.99; substantial: > 0.95–0.99; moderate: 0.90–0.95; poor: < 0.90) [17]. The results are presented as mean ± SD or mean (95% limits of agreement). IBM SPSS Statistics 26 (IBM Corp, Armonk, NY, USA) was used for Bland–Altman and bootstrap percentile calculations. Finally, the LCCC was computed using the free Medcalc © software Version 17.7.2 (Medcalc software, Ostend, Belgium) (https://www.medcalc.org/).

## Results

Finally, 41 patients participated in the study. The demographic and surgical characteristics of the patients are presented in Table 1.

**Table 1.** Demographic and surgical characteristics.

|  |  |
| --- | --- |
| Age (years) | 59 [50 to 63] |
| Sex (Male) | 13 (32) |
| Body Mass Index (kg m<sup>-2</sup>) | 24 [22 to 25] |
| ASA physical status |  |
| ASA 1 | 8 (19) |
| ASA 2 | 27 (66) |
| ASA 3 | 6 (15) |
| Type of surgery |  |
| Ovary cancer | 10 (24) |
| Colon cancer | 7 (17) |
| Pseudomyxoma | 8 (20) |
| Primary peritoneal carcinoma | 3 (7) |
| Carcinomatosis | 13 (32) |
| Anesthesia Time (min) | 545 [465 to 635] |
| Operation Time (min) | 495 [405 to 595] |
| Blood loss (ml) | 600 [320 to 1150] |

Categorical data are expressed as number and percentage n (%), and quantitative data are reported as median values and [25 to 75th] percentiles.

The mean nasopharyngeal temperature (Tnaso) after anesthetic induction was 35.9 ± 0.1 °C (35.7–36.3 °C). The temperature during HIPEC was 37.1 ± 0.7 °C (35.9 °C–37.8 °C), and subsequently, it was 36.8 ± 0.4 °C (36.2–37.7 °C).

Fig 2 illustrates the mean temperatures during the operation.

**Fig 2.**
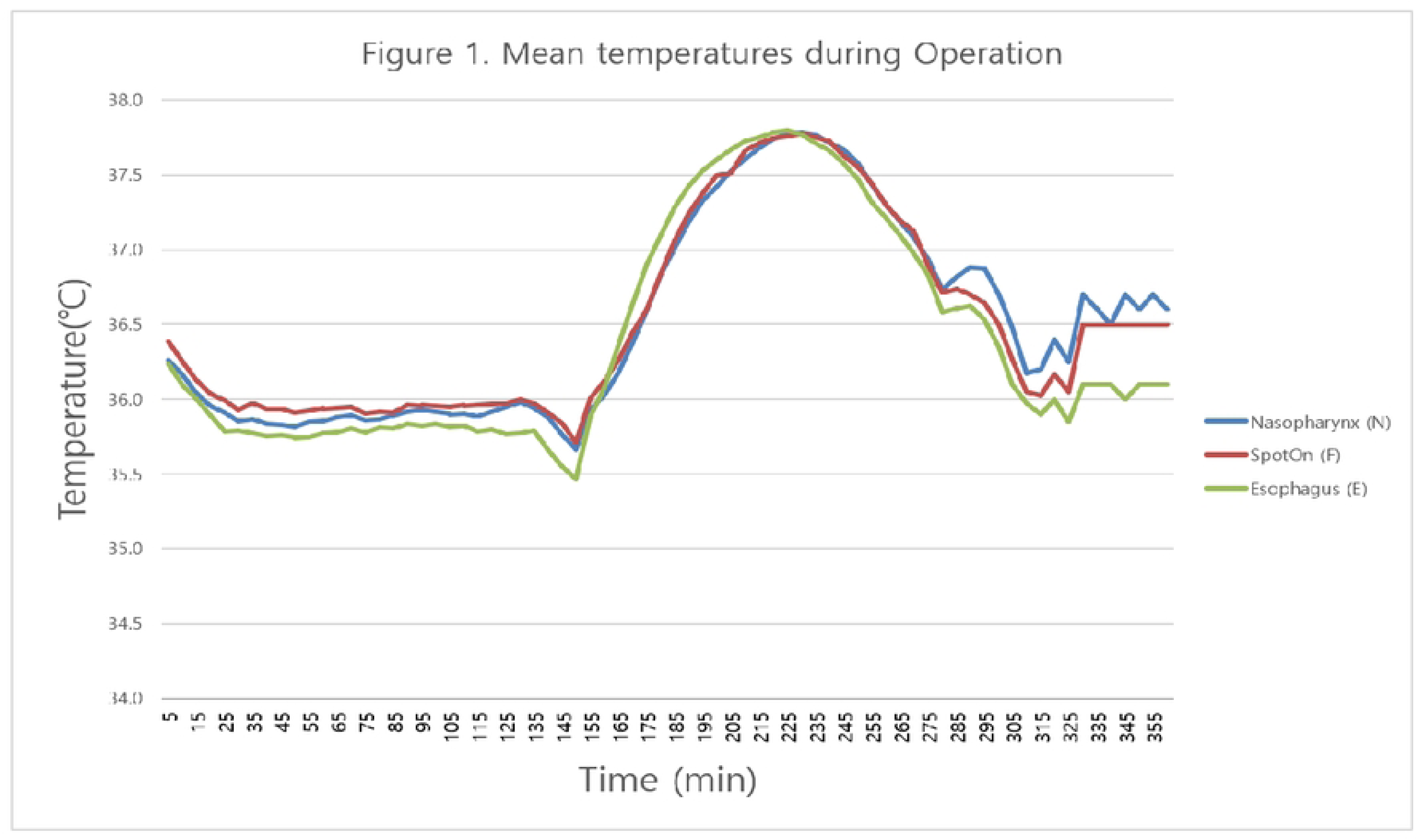
Mean temperatures during the operation.

The mean overall difference between the Tnaso and forehead (TSpotOn) temperature was -0.04 ± 0.33°C (95% limits of agreement -0.69–0.62) (Fig 3-1). The proportion of differences within 0.5°C was 0.03 (95% CI 0.01– 0.04) for nasopharyngeal reference. The LCCC was 0.94 (95% CI 0.93–0.94). (Table 2) The mean difference between Tnaso and esophageal temperature (Teso) was 0.02 ± 0.35 °C (95% limits of agreement of -0.66 – 0.71) (Fig 3-2). The proportion of differences within 0.5 °C was -0.06 (95% CI -0.08 to -0.05) for the nasopharyngeal reference. The LCCC was 0.94 (95% CI 0.93–0.94) (Table 2).

**Fig 3.**
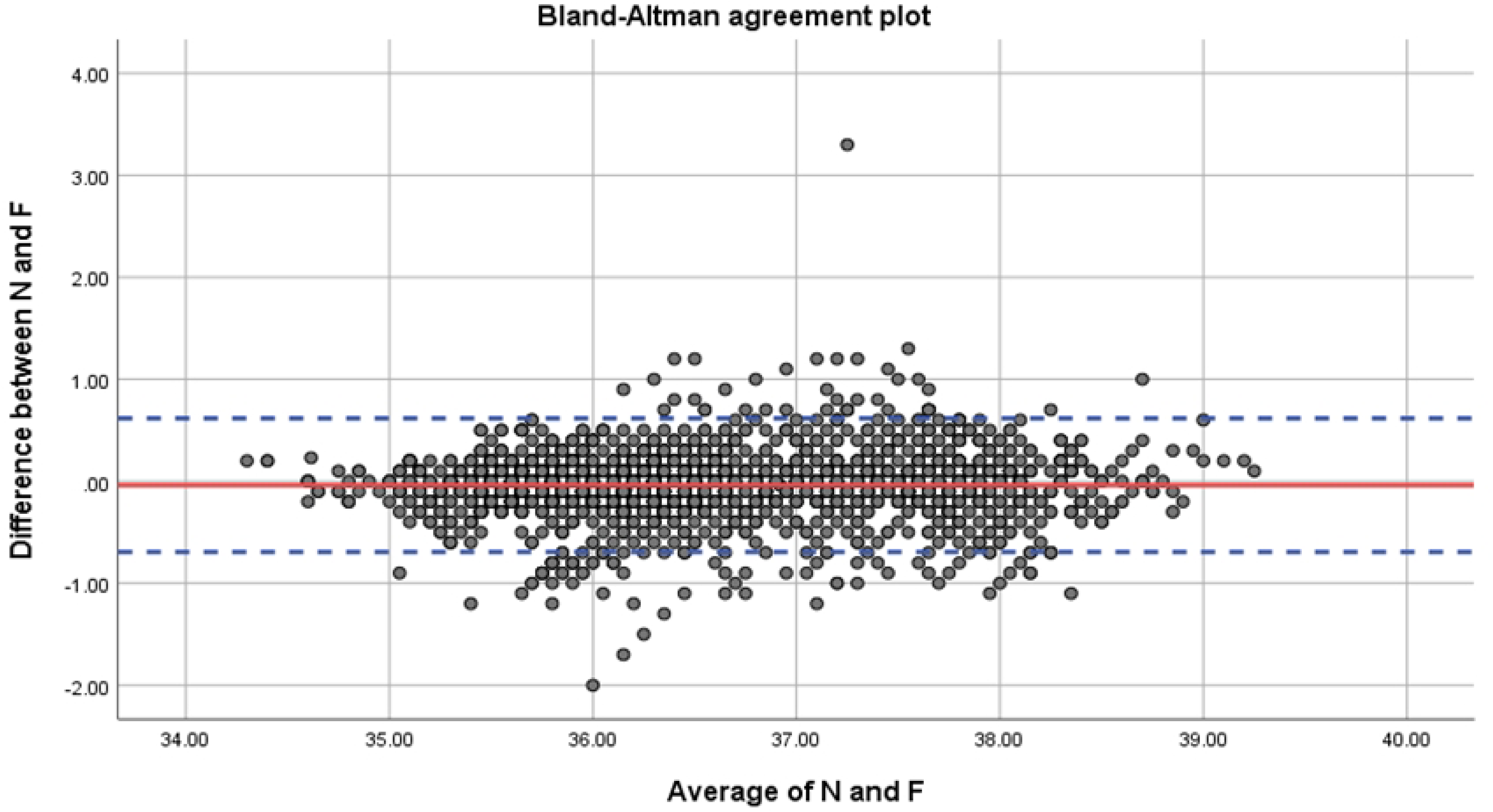
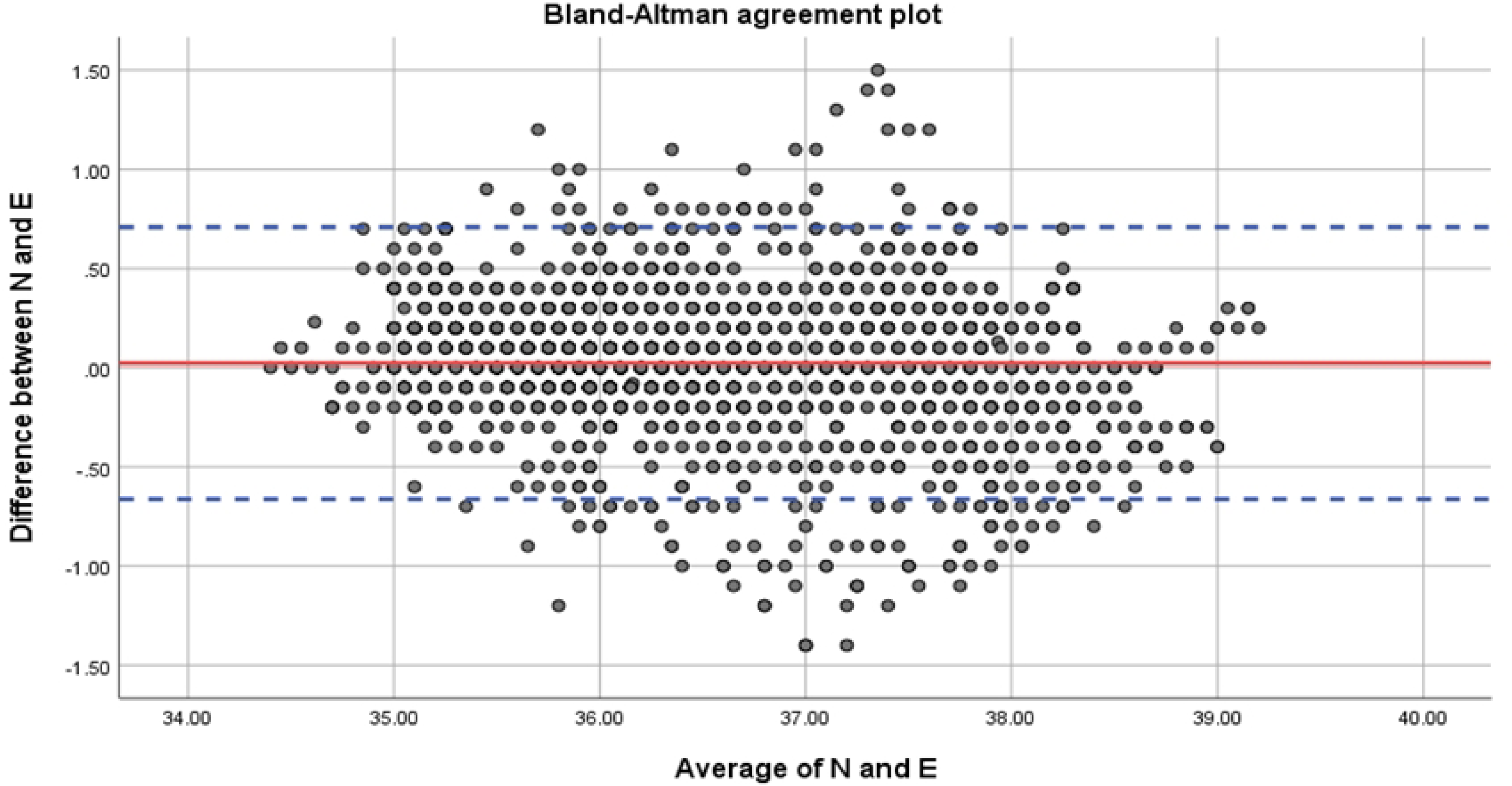
Bland–Altman plot of core temperature measurements comparing the nasopharyngeal temperature to the temperature measured by the ZHF (a) and the esophageal temperature (b). Limits of agreement (dotted lines) on the plot indicate where 95% of the differences between the two methods are expected to fall.

**Table 2.** Comparison between Tnaso and the reference methods

|  | **Mean (SD)**<br>**Temperature ( °C)** | **95% limits of agreement**<br>**(95% CI)** | **Proportion of**<br>**differences within 0.5°C**<br>**(95% CI)** | **LCCC**<br>**(95% CI)** |
| --- | --- | --- | --- | --- |
| **Overall** |  |  |  |  |
| **Teso** | 0.02 (0.35) | -0.66 – 0.71 | -0.06 (-0.08 – - 0.05) | 0.94 (0.93 – 0.94) |
| **TSpotOn** | -0.04 (0.33) | -0.69 – 0.62 | 0.03 (0.01 – 0.04) | 0.94 (0.93 – 0.94) |
| **Pre-HIPEC period** |  |  |  |  |
| **Teso** | 0.09 (0.26) | -0.42 – 0.60 | 0.00 (-0.04 – 0.03) | 0.85 (0.84 – 0.87) |
| **TSpotOn** | -0.07 (0.27) | -0.59 – 0.46 | -0.01 (-0.04 – 0.03) | 0.85 (0.83 – 0.87) |
| **HIPEC period** |  |  |  |  |
| **Teso** | -0.12 (0.39) | -0.89 – 0.65 | -0.03 (-0.06 – 0.00) | 0.90 (0.89 – 0.91) |
| **TSpotOn** | -0.03 (0.41) | -0.84 – 0.77 | 0.01 (-0.02 – 0.05) | 0.90 (0.88 – 0.91) |
| **Post-HIPEC period** |  |  |  |  |
| **Teso** | 0.15 (0.37) | -0.58 – 0.87 | -0.01 (-0.07 – 0.04) | 0.88 (0.85 – 0.90) |
| **TSpotOn** | 0.04 (0.30) | -0.55 – 0.63 | 0.03 (-0.01 – 0.07) | 0.93 (0.91 – 0.94) |

Tnaso: nasopharyngeal temperature, TSpotOn: temperatures were measured at the forehead using a SpotOn<sup>®</sup> sensor, Teso: esophageal
temperature, CI: confidence interval, SD: standard deviation, LCCC: Lin's concordance correlation coefficient

We analyzed the agreement between Tnaso and the other monitoring site temperatures (TSpotOn and Teso) separately pre-, during, and post-HIPEC. (Table 2, Figs 4-1, 2 and 5-1, 2)

**Fig 4.**
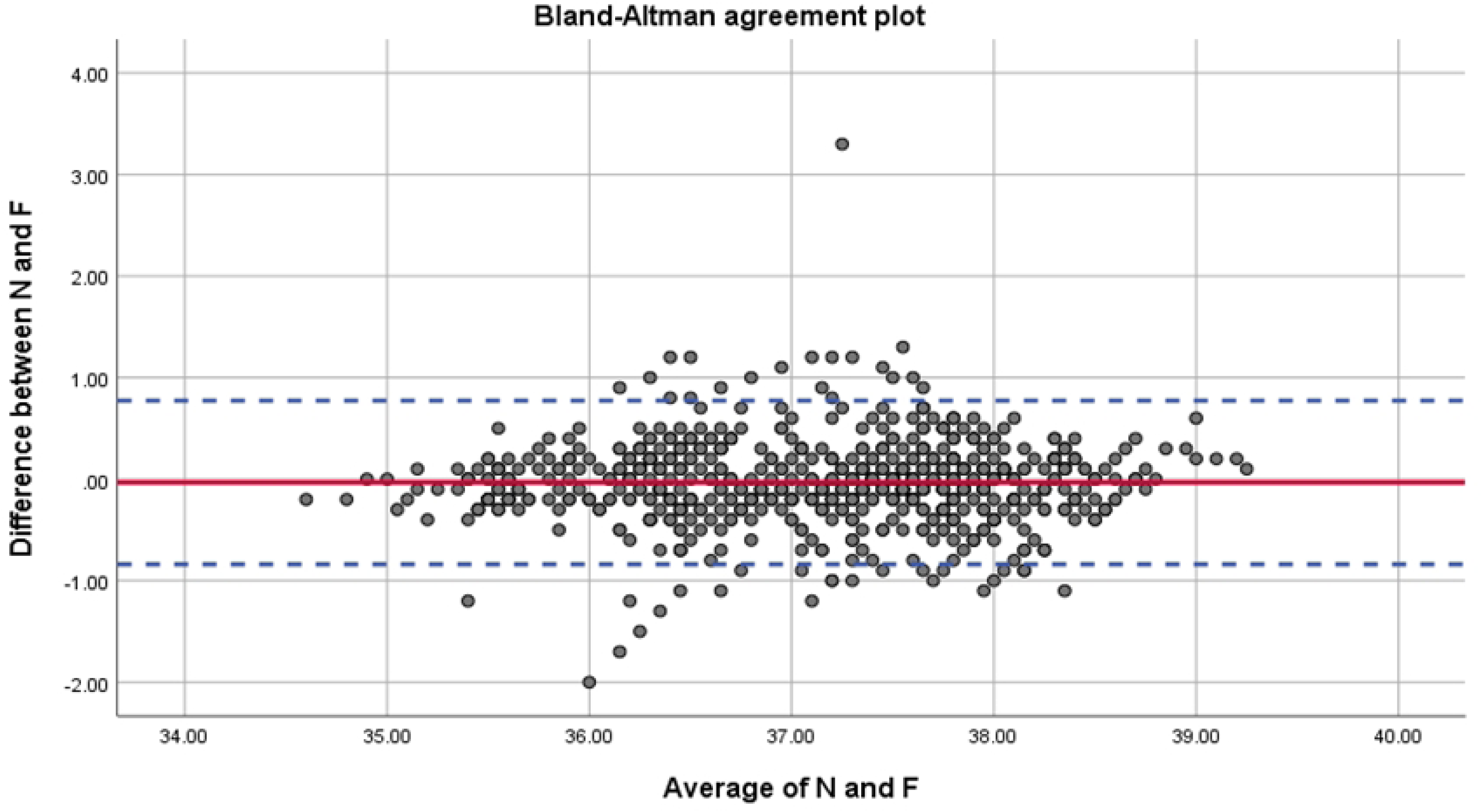
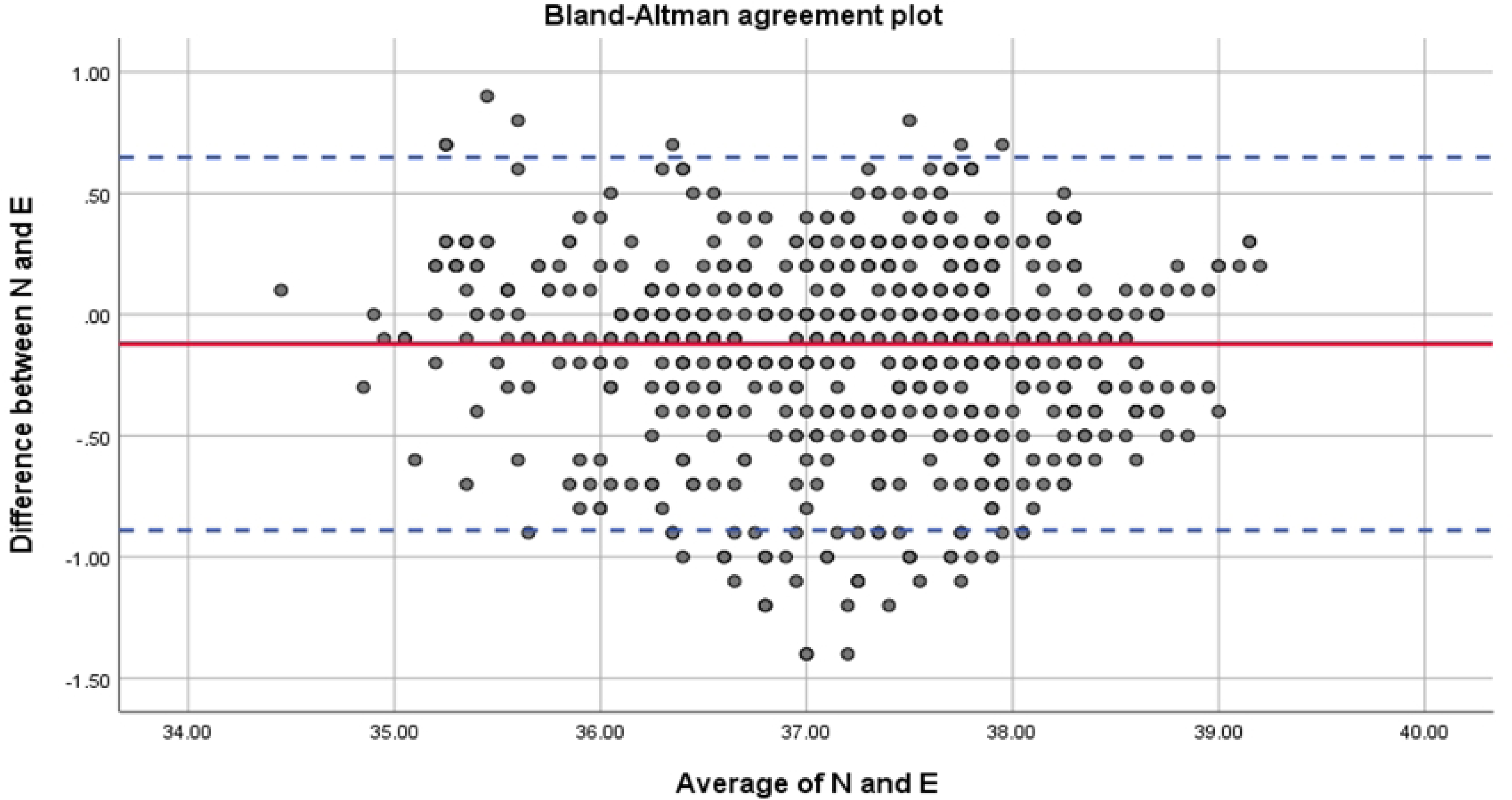
Bland–Altman plot of core temperature measurements during the HIPEC period comparing the nasopharyngeal temperature to the temperature measured by the ZHF (a) and the esophageal temperature (b). Limits of agreement (dotted lines) on the plot indicate where 95% of differences between the two methods are expected to fall.

**Fig 5.**
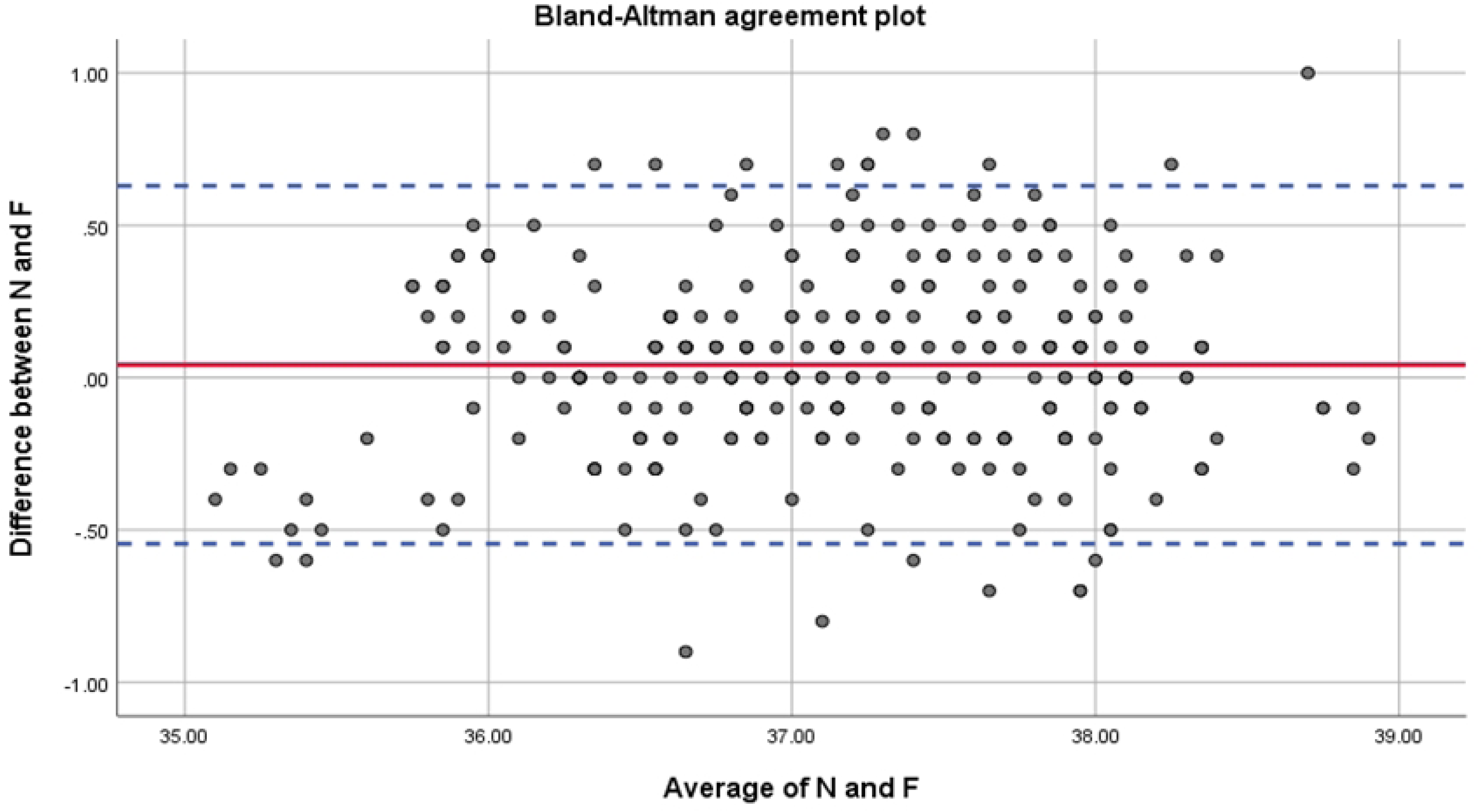
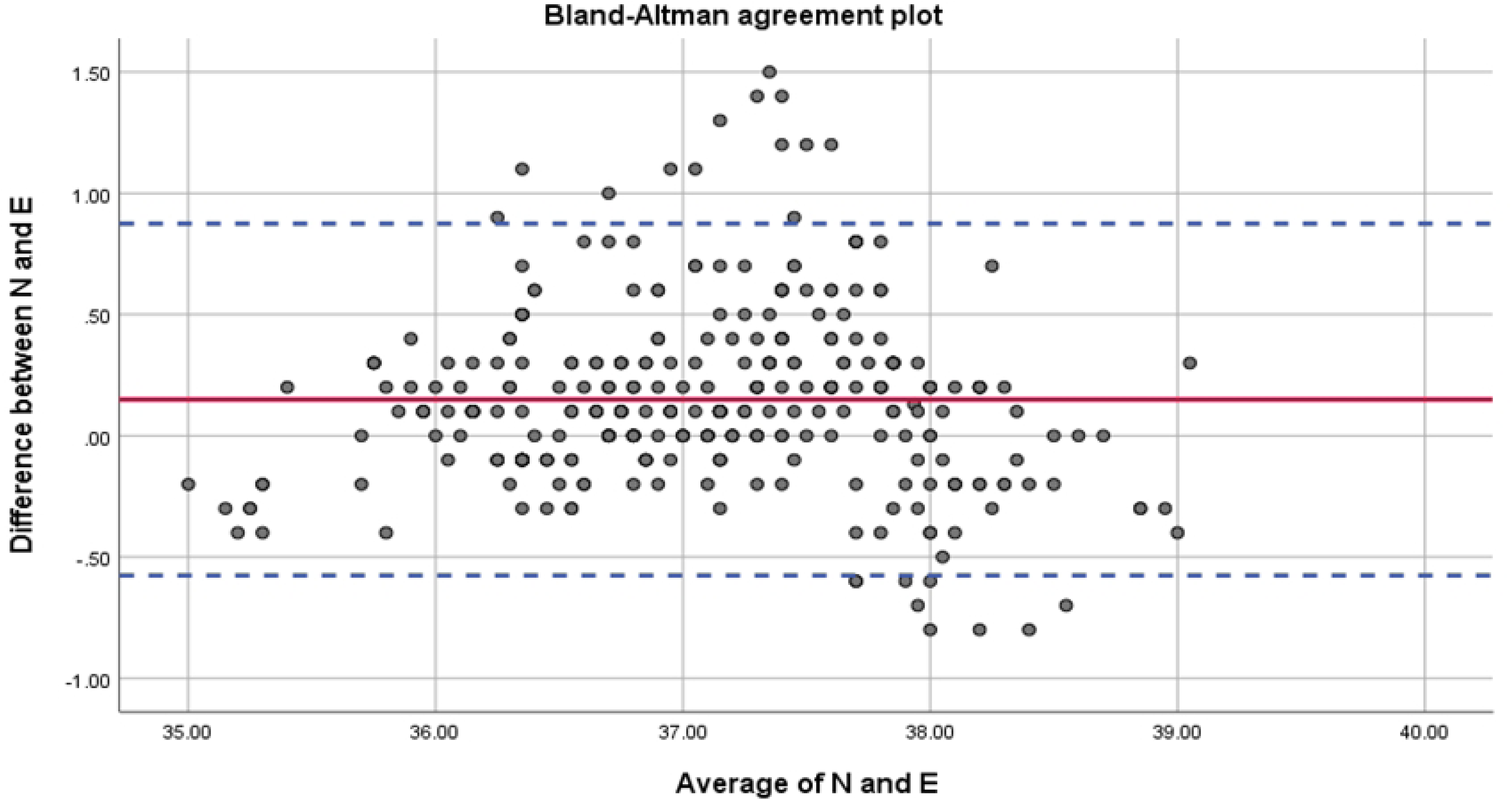
Bland–Altman plot of core temperature measurements after the HIPEC period, comparing the nasopharyngeal temperature to the temperature measured by the ZHF (a) and esophageal temperature (b). Limits of agreement (dotted lines) on the plot indicate where 95% of differences between the two methods are expected to fall.

## Discussion

Our study primarily focused on evaluating the reliability of the ZHF method during the HIPEC period. Our findings indicate a strong correlation between nasopharyngeal, ZHF, and esophageal temperatures throughout surgery.

However, in our findings, the differences in temperature pairs between Teso and other temperatures were not consistently maintained. Further, during the post-HIPEC period, rebound heating was observed across all monitoring sites, with a predominant effect in the nasopharyngeal and ZHF areas.

The nasopharynx is recommended as a reliable core temperature monitoring site in situations with wide-ranging body temperature changes, such as in cardiac surgery [18].

Additionally, since brain tissue temperature demonstrates a high correlation with nasopharyngeal temperature, monitoring brain tissue temperature can be considered a rational choice. The ZHF is an alternative non-invasive method for measuring the core temperature using a thermal insulator on the skin surface.

A recent systematic review indicated that temperature measurements from the 3M™ Bair Hugger™ Temperature Monitoring System could deviate by approximately 1 °C from the core temperature [19]. Nevertheless, our findings demonstrated a good correlation between nasopharyngeal temperature and temperature measured by the ZHF during periods of rapid core temperature changes. Esophageal temperature also showed a good correlation with nasopharyngeal temperature (LCCC 0.90, max 39.2 °C). Furthermore, throughout the pre- and post-HIPEC periods, TSpotOn and Teso exhibited LCCC values exceeding 0.8 when compared to Tnaso. During hyperthermic intraperitoneal perfusate indwelling, the nasopharyngeal core temperature increases to a mean of 37.8 °C; in our data, it rose to a maximum of 39.8 °C.

Few studies have focused on the reliability of the ZHF method during HIPEC. Boisson et al. [13] analyzed subgroup data in their observational study to assess the reliability of the ZHF and reported increasing limits of agreements during the period of rapid change in the core temperature caused by HIPEC reaching 0.6±1.8 °C, with absolute differences in temperature pairs ≤0.5 °C in less than 40% of cases. However, in our results, the mean difference compared with Teso and Tnaso, respectively, was -0.09 ± 0.46 °C, and 0.03 ± 0.41 °C, respectively, with absolute differences in temperature pairs over 0.5°C in less than 6 %. Nevertheless, in our study, 14% of pairs showed absolute differences in temperature pairs over 0.5 °C between Tnaso and Teso.

In our study, temperature data from the esophagus reflected the temperature of the abdominal cavity. During cytoreductive surgery, the overall Teso was lower than Tnaso and TSpotOn, whereas during HIPEC, Teso showed a higher value than the others. This may be explained by adjunctive tissue conduction, blood, and enthalpy of vaporization of the open abdominal cavity. The differences in temperature pairs between Teso and other temperatures were not consistently maintained; they were positive in the pre- and post-HIPEC periods but negative during HIPEC.

Interestingly, after the post-HIPEC period, the temperatures rapidly decreased, followed by a noticeable rebound increase, particularly in Tnaso and TSpotOn (Fig 2). The maximum difference between Teso and the other temperatures was 1.5°C for both Tnaso and TSpotOn. Rebound temperature changes observed in cardiac surgical patients can lead to increased cardiovascular load and collapse during rewarming, as peripheral vasodilation may cause after-drop cooling of the core temperature due to sudden return of cold blood from the extremities [20]. While no research on after-rise increase in the core temperature was found, a rebound increase in the core temperature after HIPEC was also observed in secondary cytoreductive surgery and the HIPEC study [21], possibly explaining the reverse rebound cooling process.

These discrepancies in esophageal temperature require attention during intraoperative clinical patient management. Determining the timing of cooling or heating based on the site temperature is critical, as our results indicate that Tnaso and TSpotOn were maintained at over 37 °C even after Teso dropped during the post-HIPEC period and cytoreductive surgery.

Therefore, anesthesiologists should select the core temperature monitoring site, especially for HIPEC procedures performed during laparotomy, considering the target organ.

The limitations of our study include not measuring the pulmonary artery temperature as a standard core temperature and focusing solely on cytoreductive surgery with HIPEC under laparotomy. During laparotomy, the exposed bowel and visceral heat loss through evaporation can cause exaggerated temperature discrepancies throughout the entire operative period.

In conclusion, our findings indicate a strong correlation between nasopharyngeal, ZHF, and esophageal temperatures throughout the duration of cytoreductive surgery with HIPEC. However, a considerable number of monitoring sites were required. Notably, the temperature readings in the esophagus did not match those observed in the nasopharynx and the ZHF at the forehead. Additionally, during the post-HIPEC period, rebound heating was observed across all monitoring sites, with a predominant effect in the nasopharyngeal and ZHF areas.

## Data Availability

All data files are available from the Department of Anesthesiology and Pain Medicine, CHA Bundang Medical Centervdatabase (accession via 0000-0002-2990-528X).

## Acknowledgments

Dongwook Kim assisted with the statistical analysis for this study.

## Authors’ contributions

ASW: Study design, patient recruitment, data collection, data analysis, drawing up the first draft of the paper and correspondence; JWK: statistical management, drawing up the first draft of the paper; KJH: patient recruitment, data collection, statistical management; SO: patient recruitment, and data collection. All authors have read and approved the final manuscript.

